# Acupuncture with or without acupoint application for primary dysmenorrhea: protocol for a systematic review and meta-analysis

**DOI:** 10.1101/2022.03.22.22272750

**Authors:** Xuewei Zhao, Yu Tian, Jinying Zhao, Hailin Jiang, Jiabao Sun, Xiaoyu Zhi, Baiyan Liu, Wu Liu, Yanze Liu, Tie Li, Fuchun Wang

## Abstract

**Introduction:** Dysmenorrhea is the most common gynaecological symptom reported by women. Ninety percent of women presenting for primary care experience some menstrual pain. Population surveys suggest that, although prevalence rates vary considerably by geographical location, complaints of dysmenorrhea are widespread in diverse populations. This study will evaluate the results of randomized controlled trials to determine the efficacy and safety of acupuncture with or without acupoint application for the treatment of dysmenorrhea.

**Methods:** Eight databases, including China National Knowledge Infrastructure, Chinese Scientific Journal Database, Cochrane Central Register of Controlled Trials, Embase, MEDLINE, PubMed, Wanfang Database, and Web of Science, will be searched using English and Chinese search strategies. In addition, manual retrieval of research papers, conference papers, ongoing experiments, and internal reports, among others, will supplement electronic retrieval. All eligible studies published on or before December 12, 2021 will be selected. To enhance the effectiveness of the study, only clinical randomized controlled trials related to the use of acupuncture with or without acupoint application for the treatment of dysmenorrhea will be included.

**Analysis:** The Visual Analog Scale will be the primary outcome measure, whereas the McGill pain questionnaire, SF-36 Scale, Self-Rating Anxiety, and Self-Rating Depression scale will be the secondary outcomes. Side effects and adverse events will be included as safety evaluations. To ensure the quality of the systematic evaluation, study selection, data extraction, and quality assessment will be independently performed by two authors, and a third author will resolve any disagreement.

**Ethics and dissemination:** This systematic review will evaluate the efficacy and safety of acupuncture with or without acupoint application for the treatment of dysmenorrhea. Since all included data will be obtained from published articles, it does not require ethical approval and will be published in a peer-reviewed journal.

**INPLASY registration number:** INPLASY202230051.

## Introduction

Dysmenorrhea, defined as painful menstrual cramps of uterine origin, is the most common gynecological condition among women of reproductive age[1]. Dysmenorrhea is classified as primary or secondary based on whether or not an underlying etiology is identified.[2] Primary dysmenorrhea(PD)is pain with menses for which there is no underlying abnormality, whereas secondary dysmenorrhea is pain associated with conditions such as endometriosis, pelvic inflammatory disease, leiomyomas, and interstitial cystitis.[3] Treatment of secondary dysmenorrhea focuses on the causative pelvic pathology or medical condition. Primary dysmenorrhea accounts for the majority of painful menses in ovulatory women.

Reported prevalence varies widely, ranging from 17% to as high as 90%[4.5]. Ag e is inversely related to menstrual pain,13 with symptoms being more pronounced in adolescents.[6.7.8] There is some evidence that parous women tend to have less dysmenorrhea. Some women experience relatively minimal pain, whereas others are significantly limited in their ability to function during their menses. Up to 15% of women with dysmenorrhea experience symptoms of sufficient severity to cause absenteeism fro m work, school, and other activities.[5.9] Generally, hormonal contraception and NSAIDs are considered first-line therapy for primary dysmenorrhea. Ibuprofen and naproxen are generally affordable and well tolerated, and both can be titrated to balance efficacy with side effects. Contraindications to the therapy, frequency of administration, orprohibitive side effects may render this option unacceptable. Non-pharmacological complementary therapies may be associated with new forms of drug administration and thus also reduce the level of drug exposure and its side effects in order to potentiate relief of the symptoms of primary dysmenorrhea.[10] The application of acupoint stimulation (acupressure and acupuncture), alone or in combination with other therapies, has been the subject of active investigation in recent years.[11] These are commonly used by women in an effort to relieve dysmenorrhea.[12]

Currently, the therapy levels of the available evidence-based research on acupuncture for the treatment of PD. This is because the existing studies have limitations such as small sample size, loosely implemented randomization, different degrees of blinding, use of nonstandard intervention measures in the control groups, and lack of evidence from high-quality randomized controlled trials (RCTs). Therefore, the aim of this study is to analyze the results of RCTs to ascertain the efficacy of acupuncture for the treatment of PD. This will allow for the provision of reliable evidence-based clarification of the efficacy of acupuncture for the treatment of PD.

The proposed date for the completion of this study is December 12, 2021.

## Materials and methods

This protocol is based on the Preferred Reporting Items for Systematic Reviews and Meta-Analyses Protocols (PRISMA-P) guidelines[13] and the corresponding checklist. The systematic review protocol is registered in the INPLASY International Registry of Systematic Reviews(ID: INPLASY202230051; https://inplasy.com/?s=INPLASY202230051; DOI: 10.37766/inplasy2022.3.0051).

### Inclusion and exclusion criteria

#### Types of participants

Patients with abdominal pain before or after menstruation or during menstruation, mainly concentrated in the lower abdomen, and with none of the other symptoms, including headache, dizziness, nausea and vomiting, diarrhea, waist and leg pain[11]. primary dysmenorrhea is characterized by crampy, suprapubic pain that begins between a few hours before and a few hours after the onset of the menstrual bleeding. Symptoms peak with maximum blood flow. regardless of their sex and nationality. Exclusion criteria will include1) Secondary dysmenorrheainclude Endometriosis, Aden omyosis, Uterine myomas, Cervical stenosis, Obstructive lesions of the genital tract. 2) Other causes of menstrual pain may include the following: Pelvic inflammatory disease, Pelvic adhesions, Irritable bowel syndrome, Inflammatory bowel disease, Interstitial cystitis, Mood disorders, Myofascial pain.

#### Types of interventions

We will include studies in which the intervention group received acupuncture alone or in combination with acupoint application, and in which the control group received without acupuncture treatment, sham acupuncture group, or conventional treatment.

#### Types of outcome measures

The primary outcome measure will be the extent of pain in the lower abdomen measured by visual analog scale (VAS), and relief from symptom s.The additional outcomes will be quality of life(QoL), and adverse events.

Different acupuncture manipulation will be analyzed in subgroup analyses.

#### Types of studies

Randomized controlled clinical trials and quasi-RCTs will be includeed.

### Data sources

A structured and systematic literature search for eligible and relevant articles published on or before January 15, 2022 will be conducted. The following databases will be searched: China National Knowledge Infrastructure, Chinese Scientific Journal Database(VIP), Embase, MEDLINE, PubMed, Wanfang Database, and Web of Science. Selected studies will be published in either Chinese or English. The search terms include “primary dysmenorrhea”, “acupuncture”, “acupuncture therapy”, “acupoint application”, “randomized controlled trial”, “acupoint”, “clinical RCT”, “trial”, “groups”.

### Searching other resources

TA manual search will mainly be used in searching for relevant studies. Details of the selection process are shown in a flow chart and the screening process is summarized in a flow diagram (Fig1)

### Search strategy

The search strategy will be based on the Cochrane handbook guidelines (5.1.0) and will include keywords, such as “acupuncture”, “acupoint application”, “primary dysmenorrhea” o r “acupoint”, “randomized controlled trial”, “clinical RCT”, “trial”, “groups”. Subsequent searches will involve the use of Medical Subject Headings terms headings, in addition to keywords from the initial retrieval. Additional article searches will involve a review of the reference lists of relevant research articles. As an example, the search strategy for PubMed is summarized in Table 1

**Table 1.** Search strategy for PubMed

| Number | Search terms |
| --- | --- |
| 1 | acupuncture. ti, mesh. |
| 2 | Acupuncture Analgesia. ti, ab. |
| 3 | acupuncture therapy. ti, ab. |
| 4 | acupoint application. ti, mesh. |
| 5 | Acupuncture point. ti, ab. |
| 6 | or 1-5 |
| 7 | dysmenorrhea. ti, mesh |
| 8 | primary dysmenorrhea. ti, ab. |
| 9 | menstrual pain. ti, ab. |
| 10 | painful menstruation. ti, ab |
| 11 | or 7-10 |
| 12 | randomized controlled trial. pt. |
| 13 | controlled clinical trial. pt. |
| 14 | randomized. ab. |
| 15 | Randomly. ab. |
| 16 | trial. ab. |
| 17 | or 12-16 |
| 18 | 6 and 11 and 17 |

### Selection of studies

Two researchers (JBS and XYZ) will independently select the eligible literature according tothe inclusion and exclusion criteria after reading their titles and abstracts. Subsequently, the full texts of the papers will be read and uncontrolled research, nonrandomized studies, and studies with inconsistent evaluation criteria or similar data will be excluded. If any differences occur during the screening, the third author (HLJ) would intervene.

### Data extraction and management

Two researchers (JBS and XYZ) will use a predesigned data extraction table to extract the data of the included studies. The extracted data will include author, year, sample size, course ofMtreatment, intervention measures, outcome indicators, adverse reactions, etc. The study selection procedure will be performed according to the PRISMA guidelines, which a represented in the flow diagram (Fig 1).

**Figure 1.**
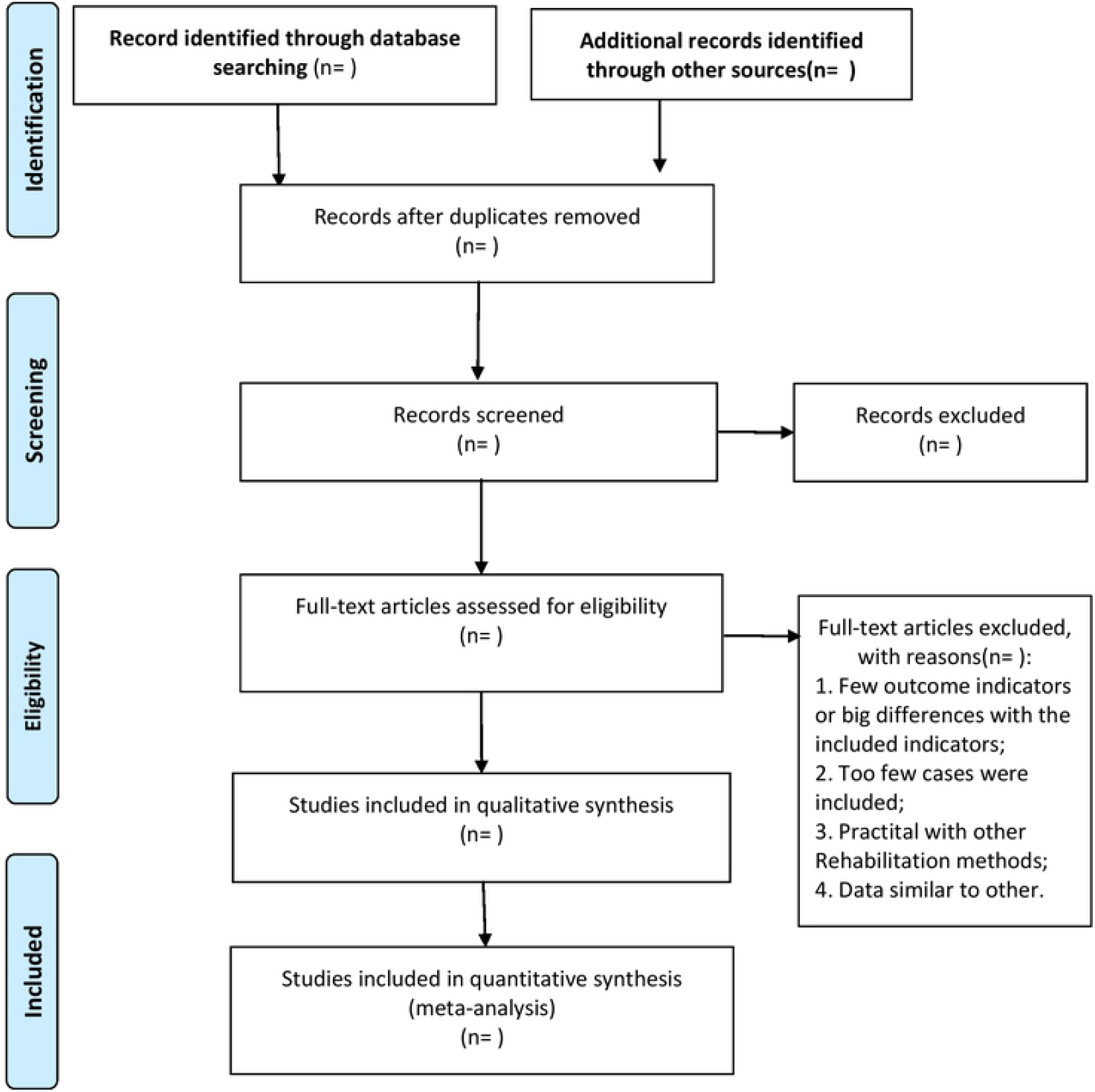
PRISMA flow diagram of study and exclusion.

### Statistical analysis

We will use SAS (SAS Institute, Cary, NC) and Stata (StataCorp, College Station, TX) to analyze the standard deviation, standard error and mean of each group. When we encounter literature with missing data, we will try to contact the author. If we are unable to get full data, then that study will not be included. We will use the Review Manager 5.4 softw are provided by the Cochrane Collaborative Network for statistical analysis [14]. For continuous variables, the means and standard deviations of each study will be obtained and pooled as a mean difference or a standardized mean difference with a 95% confidence interval. The statistical heterogeneity of the included clinical RCTs will be analyzed. The I2 test will be used to test for heterogeneity. When I2 is <50% or P>0.05, it indicates that there is no statistical heterogeneity between studies [15]. A fixed-effects model will be selected to combine the effect amount; otherwise, a random effects model will be adopted.

### Methodological assessment of quality

The qualities of the included studies will be evaluated using the risk of bias table proposed by the Cochrane Collaborative Network [15]. The risk table includes six items: random sequence generation mode, whether to use allocation concealment, whether to blind the subjects and intervention providers, whether to blind the result evaluators, whether the result data are complete, whether to select the result report, and other bias sources. The criteria used to assess the risk of bias are “low risk”, “high risk”, and “unclear” [15]. Two evaluators will independently evaluate the methodological qualities of the studies. In cases of disagreement, the third author would intervene.

### Assessment of heterogeneity

If there is no significant heterogeneity (I2<50%)between studies, the fixed-effects model will be used for evaluation. If there is significant heterogeneity (I2>50%), the random-effects model will be used for evaluation [15]. Sensitivity analysis or subgroup analysis will then be conducted as required to explain the heterogeneity.

### Subgroup analysis

If possible, subgroups will be analyzed according to different acupuncture manipulation.

### Sensitivity analysis

When possible, we will perform sensitivity analysis to explore the effects of a trial’s risk of bias on primary outcomes. Lower quality trials will be excluded from these analyses and the meta-analyses will be repeated according to sample size and insufficient data to assess quality and robustness when significant statistical heterogeneity arises.

### Assessment of publication bias

If more than 10 trials meet the study criteria, we will use Review Manager 5.4 software t o draw and analyze a funnel chart and use the funnel chart to evaluate the potential publication bias.

### Grading the quality of evidence

The Grading of Recommendations Assessment, Development and Evaluation (GRADE) app roach [16] is recommended for the analysis of the level of evidence.

## Discussion

Many studies in the current literature have shown that a range of risk factors might be associated with PD, including biological, psychological, social, and lifestyle factors. Biological factors might include earlier age at menarche, heavier menstrual flow, and family history of dysmenorrhea[17.18]; psychological factors include stress, anxiety, and depression[19]; social factors include a lower level of social support[20]; and lifestyle factors include cigarette smoking and irregular diet.[5] As a debilitating condition for many women, dysmenorrhea is one of the leading causes of absenteeism from school or work, which has a negative effect on quality of life, daily living, work productivity, and academic activities.[21.22] Dysmenorrhea is an emerging serious gynecological and public health problem among university students, which highlights the need for better intervention and prevention of this condition.

### Acupuncture with acupoint application has become an important method for primary dysmenorrhea

Acupuncture for the treatment of dysmenorrhea has unique advantages[23]. With the increasingly widespread use of acupuncture globally, the mechanism of acupuncture in primary dysmenorrhea has become a research hotspot. Mechanism of acupuncture treatments manifested by different brain responses.[24.25] Acupuncture treatment has been widely accepted because of its simplicity, convenience, fewer side effects, and exact curative effect.

### Clinical effect of acupuncture in the treatment of dysmenorrhea is better than that

There are many reports on the treatment of dysmenirrhea by acupuncture with acupoint application and acupuncture with moxibustion. While the moxibustion procedure is simple th anacupoint application, we found that the effect of acupuncture with acupoint application is better through clinical research. Acupuncture combined with acupoint application can relieve abdominal pain symptoms and reduce pain more than acupuncture combined with moxibustion[26]. Acupuncture combined with acupoint application therapy is more convenient and cost-effective.

### VAS and other outcome indicators are closely related to dysmenorrhea

VAS used to assess pain intensity in diverse clinical settings.[27] A preponderance of evidence demonstrates that the 100 mm visual analog scale (VAS) is by far the most frequently used assessment instrument to evaluate analgesic effects of various therapies.[28.29] In addition, the McGill and quality of life are other scales for assessing pain and often used in the assessment of dysmenorrhea.[30.31] Combined with these evaluation methods, they can be evaluated from multiple angles and play a complementary role, making the evaluation results more reliable. The mental state of dysmenorrhea is also very important for the recovery of dysfunction. There is currently no commonly used scale to evaluate the mental state of stroke patients in the literature.

There are currently no data on the systematic reviews or meta-analyses of the therapeutic effect of acupuncture wiht or without acupoint application on dysmenorrhea. Therefore, we plan to conduct a systematic review and meta-analysis to provide high-quality evidence of the efficacy of acupuncture with acupoint application for dysmenorrhea.

### Limitations

As acupuncture with acupont application is a holistic method and follows the concept of “treatment based on syndrome differentiation,” different patients and different acupuncturists will produce different therapeutic effects, leading to a certain level of heterogeneity. In addition, different types of acupuncture, including reinforcing and reducing, flat reinforcing and reducing, and other manipulations, also produce different therapeutic effects. Second, the following drawbacks cannot be avoided during literature collection: the quality of literature not being high enough, the exclusion of studies not published in Chinese or English, and the sample size of the included literature not being large enough.

## Data Availability

Eight databases, including China National Knowledge Infrastructure, Chinese Scientific Journal Database, Cochrane Central Register of Controlled Trials, Embase, MEDLINE, PubMed, Wanfang Database, and Web of Science

https://www.cnki.net/

https://pubmed.ncbi.nlm.nih.gov/

## Supporting information

S1 Checklist. PRISMA-P (Preferred Reporting Items for Systematic review and Meta-Analysis Protocols) 2015 checklist: Recommended items to address in a systematic review Protocol^*^. (DOC)

## Author Contributions

Conceptualization: Xuewei Zhao.

Data curation: Jiaobao Sun, Xiaoyu Zhi.

Formal analysis: Hailin Jiang.

Funding acquisition: Fuchun Wang.

Methodology: Yu Tian, Jinying Zhao.

Software: Baiyan Liu, Wu Liu, Yanze Liu.

Supervision: Tie Li.

Writing – original draft: Xuewei Zhao.

Writing – review & editing: Xuewei Zhao, Tie Li, Fuchun Wang.

